# Quantitative Insights into CAR-T Cell Therapy: The Interplay of Dose, Dosing Regimen, Proliferation and Tumour Elimination

**DOI:** 10.1101/2025.09.02.25334722

**Authors:** Shilian Xu, Maoxuan Liu, Jiaru Yang

## Abstract

Chimeric antigen receptor (CAR)-T cell therapy marks a significant advance for patients with B-cell acute lymphoblastic leukemia. Clinically, CAR-T cell doses are precisely calculated based on the patient’s weight, especially for paediatric patients. Patients with higher tumour burdens at the start of CAR-T therapy are less likely to both attain and maintain a deep response compared to those with lower tumour burdens. To quantitatively investigate how CAR-T cell dose, dosing regimen and tumour burden jointly determine therapy outcomes, we developed a family of mathematical models. We first analysed flow cytometry-based killing assay data testing RAJI-19 cells against CAR-T cells, and found that CAR-T cell lysing efficiency increases but saturates with further increases of both RAJI-19 cells and CAR-T cells. This interaction leads to bistable RAJI-19 cell kinetics; specifically, low tumour burdens are effectively inhibited, while high tumour burdens remain refractory. Our models predict that high CAR-T cell proliferation inhibit RAJI-19 cell kinetics independent of dosing regimens. However, with fixed total dose, single-dose infusion provides superior outcomes when proliferation is low. The predicted bistable CAR-T cell concentration interval matches with observed post-infusion CAR-T cell concentrations. Our findings offer a potential mechanistic explanation for observed variations in therapy outcomes and inform personalized CAR-T cell therapy.

## Introduction

Chimeric antigen receptor (CAR) T cell therapy, involving the genetic modification of a patient’s own T cells to target tumour-associated antigens, has revolutionized clinical outcomes for patient with advanced acute lymphocytic leukemia and B-cell lymphomas [1]. CAR-T cell therapy is now approved by the FDA (US Food and Drug Administration) to treat several blood cancers, including B-cell acute lymphoblastic leukemia (B-ALL), chronic lymphocytic leukaemia (CLL), large B-cell lymphoma (LBCL) and multiple myeloma (MM), with remission rates of 76% in B-ALL [2], 89% in MM [3] and 29% in CLL [4].

Both disease-specific factors and therapy-specific factors collectively determine CAR-T cell therapy outcomes. These factors include tumour burden [4, 5], CAR design [6, 7], CAR-T cell dose [2, 8], proliferation and persistence [9, 10], all of which exhibit significant variability among patients. Critically, among B-ALL patients, those with higher tumour burdens at the start of CAR-T treatment are less likely to both attain and maintain a deep response compared to those with lower tumour burdens; specifically, the probability of durable response decreases with increased tumour burden [5, 11]. Despite this, current weight-based CAR-T cell dosing strategies (e.g., viable CAR-T cells per kg recipient body weight or per m^2^ recipient body surface area [8, 12]) are largely empirical. They often fail to fully account for the complex interactions between tumour and CAR-T cells, which fundamentally dictate CAR-T cell persistence, proliferation, and overall efficacy against tumour burden.

High tumour burden and its associated impaired immunity play essential roles in CAR-T cell therapy failure [13]. Firstly, large tumour burdens create a more significant immunosuppressive environment, hindering the host’s natural immune responses and its ability to effectively mount immunotherapy-induced responses [14]. Second, CAR-T cells harvested from patients with high tumour burdens can show impaired fitness, leading to a decreased ability to adequately proliferate and control tumour growth [15, 16]. Finally, the severity of CAR-T cell-related systemic toxicities, like cytokine release syndrome (CRS) and immune effector cell– associated neurotoxicity syndrome (ICANS), is strongly linked to the tumour burden [5, 13, 17].

Mathematical models have significantly advanced our understanding of *in vivo* and *in vitro* tumour kinetics in leukaemia [18-22], and have been instrumental in quantifying the role of immunotherapy [19, 23-27], particularly CAR-T cell dose, dosing, tumour burden and exhaustion. Flow cytometry-based killing assays for allowed the estimation of tumour cell and viable remaining CAR-T cell concentration before and over co-culture incubation, and determined the percentage of live and dead target cells under different conditions [28, 29]. A variety of mathematical models were proposed to understand killing assay datasets. For example, Prativa Sahoo and her colleagues use law of mass action to delineate real-time cytotoxicity assay datasets [18]. Kirouac et al utilized Hill function with respect to CAR-T cell concentration to describe CAR-T cell lysing [24, 30], suggesting that CD19+ cell kinetics depend on magnitude of CAR-T cells rather than CD19+ cell. Chauhury et al [31] reviewed different CAR-T cell lysing formula by CAR-T cells, including mass action functional 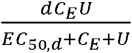, where *C*_*E*_ represents CAR-T cell concentration, *U* represents tumour cell concentration, *d* represents CAR-T cell lysis rate, and *EC*_50,*d*_ represents the rising section of the sigmoid curve. However, this formula fails to consider that tumour cell and CAR-T cell may play different roles to control saturation effect. Clinically, at least therapy outcomes are observed in CAR-T cell therapy outcomes on B-ALL: complete response (CR), partial response (PR) and stable disease (SD) [2, 32, 33]. It remains unclear how mathematical models can comprehensively capture CAR-T cell dose, dosing regimen, and tumour burden to determine various therapy outcomes.

AUTO1 is a fast off-rate CD19-targeting CAR with a 41BBz endodomain, which exhibits low immunotoxicity and side effects [28, 29]. Clinical trials (CARPALL (NCT02443831) and ALLCAR19 (NCT02935257)) have evaluated AUTO1’s performance on paediatric and adult B-ALL [34, 35]. Despite excellent clinical response rates, a proportion of patients experienced relapse with CD19+ disease. To investigate factors contributing to this, Reference [28] explored pharmacological inhibition of the T cell signalling pathway through AKT. Mehra et al. used AUTO1 CAR-T cells manufactured at small-scale, both with AKT inhibitor VIII (VIII-CAR) and without (untreated, UT-CAR), from healthy donor T-cells and B-ALL ALLCAR19+ patients (CAR-T cell manufacture, *Method* and [28]). AKT inhibitor VIII, a cell-permeable quinoxaline compound, exhibits a high capability to limit T-cell differentiation, enhancing T-cell ‘fitness’ and improving adoptive cell therapy products [28]. Herein, we quantitatively delineate CAR-T cell lysing efficiency by analysing flow cytometry-based killing assay (FBK) data from Reference [28].

To quantitively understand how tumour burden, dosing regimen, and CAR-T cell dose determine RAJI-19 cell kinetics, we developed a family of two-dimensional mathematical models incorporating CAR-T cell-mediated tumour lysis. These models were fitted to flow cytometry-based killing assay (FBK) data of RAJI-19 cells against VIII-CAR T cell (AUTO1 manufactured with AKT inhibitor VIII) and UT-CAR T cell (AUTO1 without AKT inhibitor VIII). Our model incorporates CAR-T cell lysing efficiency that saturates with increasing concentrations of either RAJI-19 cells or CAR-T cells, a finding that effectively fit the FBK assay data for both VIII-CAR and UT-CAR products. The interaction of RAJI-19 cells and CAR-T cells leads to bistable RAJI-19 cell kinetics, where high tumour burdens are not cleared, but low tumour burdens are effectively controlled. We found that VIII-CAR T cells resulted in a narrower CAR-T cell concentration interval for RAJI-19 cell kinetic inhibition compared to UT-CAR T cells. Our model further predict that high CAR-T cell proliferation enhances tumour inhibition regardless of the dosing regimen (e.g., single-dose or split-dose infusion). However, with a fixed infusion dose, single-dose infusion suppresses RAJI-19 cell kinetics more effectively when CAR-T cell proliferation is low. The predicted bistable CAR-T cell concentration interval aligns well with observed post-infusion CAR-T cell concentrations (median:480.5, range: 39.7-11,346 cells/μL [36]; geometric mean: 1098.79 cells/μL, range: 10^2^-10^4^ cells/μL [29]). Overall, this analysis reveals a potential mechanistic explanation for observed variations in therapy outcomes and inform personalized CAR-T cell therapy.

### Preliminary background

We included three experimental groups in our study: non-transduced T cell (NT), AUTO1 manufactured with AKT inhibitor VIII (VIII-CAR), AUTO1 manufactured without AKT inhibitor VIII (untreated, UT-CAR). These were generated using T cells obtained from six healthy donors and six B-ALL patients(shown in Fig. 1A and 1B) [28]. Of the six B-ALL patients, three were in ongoing long-term remission, and three had experienced CD19+ relapse. CAR-T cells were directly tested against RAJI-19 cells following manufacture, which was known as the ‘end of manufacture’ condition (shown in Fig.1C). In parallel, to uncover more subtle functional differences, CAR-T cells were cocultured with RAJI-19 cells for seven days before initiating flowcytometry-based killing assay. This ‘stress’ model was also known as the ‘rechallenge’ CAR-T cell assay (shown in Fig.1C).

**Figure 1.**
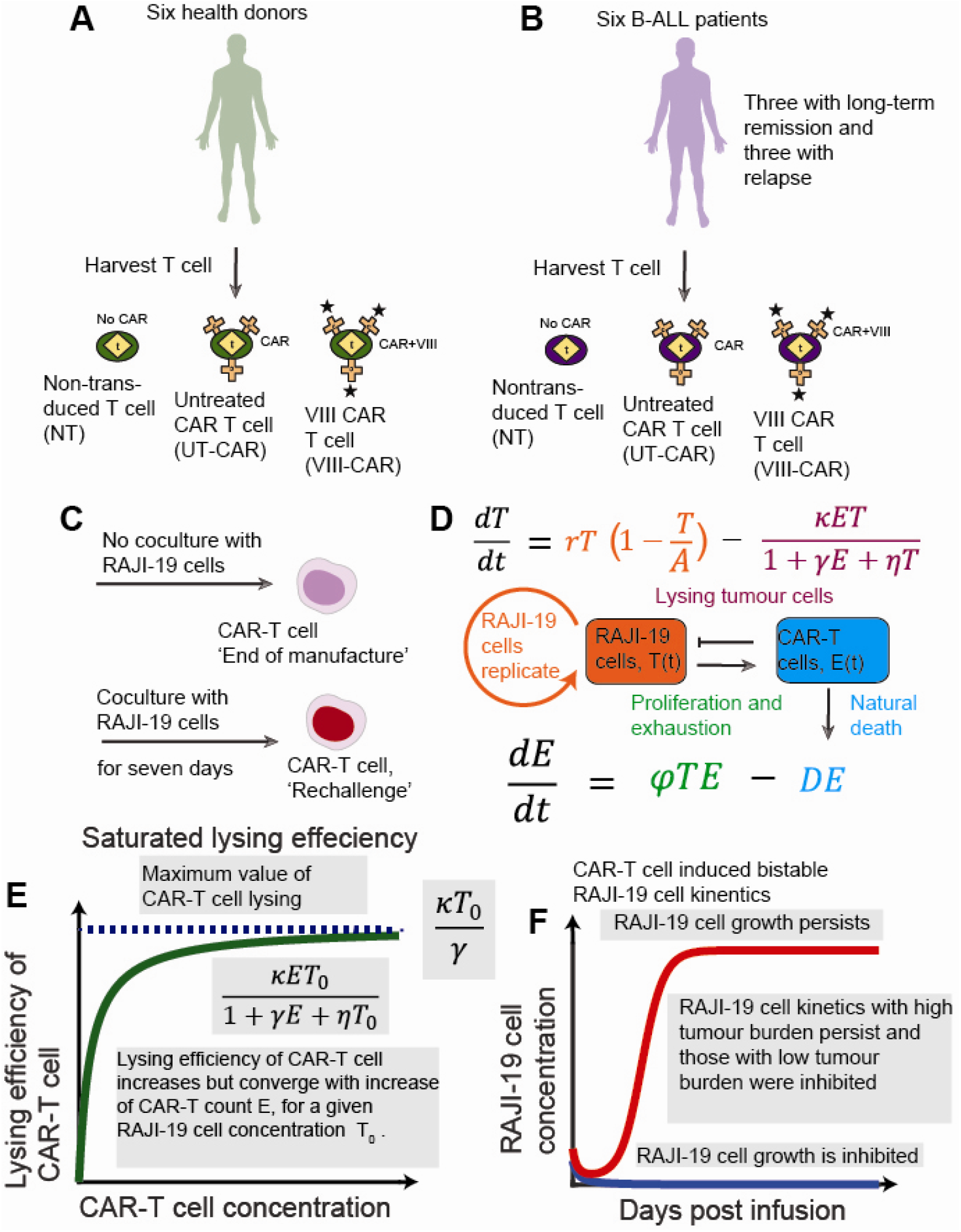
Experimental Design, Mathematical Model Framework, and Bistability Hypothesis in CAR-T Cell Therapy. **(A, B)** T cells used in this study were obtained from six healthy donors and six B-cell acute lymphoblastic leukemia (B-ALL) patients. The patient cohort included three individuals with ongoing long-term remission and three who had experienced relapse. From these sources, three experimental CAR-T cell groups were generated: non-transduced T cells (NT), AUTO1 manufactured with AKT inhibitor VIII (VIII-CAR), and AUTO1 manufactured without AKT inhibitor VIII /untreated (UT-CAR). **(C)** CAR-T cells were evaluated using two primary assay conditions: the ‘end-of-manufacture’ assay, where CAR-T cells were directly tested against RAJI-19 target cells after production, and the ‘rechallenge’ assay, where CAR-T cells were co-cultured with Raji-19 cells for seven days to induce stress before cytotoxicity testing. **(D)** Model schematic diagram illustrating the RAJI-19 cell kinetics interacting with CAR-T cells. The system is conceptually divided into two compartments: RAJI-19 cell growth follows logistic growth 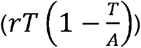, and CAR-T cells lyse RAJI-19 cells 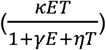. The model also accounts for Raji-19 cell proliferation and exhaustion (*φTE*), alongside their natural degradation rate (*DE*). **(E)** Illustration of saturated lysing efficiency. This refers to the phenomenon where the lysing efficiency of CAR-T cells increases but gradually approaches a maximum value as either CAR-T cell concentration or tumour cell concentration increases. **(F)** Representation of CAR-T cell-induced bistable Raji-19 cell kinetics. This hypothesis posits that within a specific range of CAR-T cell concentrations, Raji-19 cell kinetics with low tumour burdens are effectively inhibited, while those with high tumour burdens persist.

We define CAR-T cell lysing efficiency as the rate at which RAJI-19 cells are lysed, thereby quantifying anti-tumour stress exerted by CAR-T cells. Quantifying *in vivo* and *in vitro* anti-tumour pressure of CAR-T cells often necessitates mathematical modelling. The RAJI-19 cell kinetics interacting with CAR-T cells is modelled within two compartments. RAJI-19 cell growth follows logistic growth 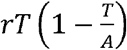 [24], and CAR-T cells lyse RAJI-19 cells. The model also assumes CAR-T cells proliferate and exhaust at rate *φ*, alongside CAR-T cell natural degradation rate *D* (shown in Fig. 1D). The specific functional form describing how CAR-T cells lyse RAJI-19 cells will be determined by fitting our proposed models to the flow cytometry-based killing assay data. Biologically, CAR-T cells do not immediately proliferate after simulation with RAJI-19 cells; namely, a three-day lag exists for CAR-T cell proliferation after initial simulation (Fig.1F, [37]). Since the flow cytometry-based killing assay co-cultures RAJI-19 cell and CAR-T cell for three days, CAR-T cell concentration decrease and CAR-T cell proliferation thus can be ignored; in this case, we used System 1 - System 4 in Methods to describe CAR-T cell and RAJI-19 cell kinetics over flow cytometry-based killing assay. To qualitatively describe CAR-T cells proliferation for long-incubation *in vivo* and *in vitro* assays, we employ System 5 to delineate the RAJI-19 cell kinetics interacting with proliferating CAR-T cells (also shown in Figure 1D) [18].

Mathematical models often use the law of mass action to describe CAR-T cell lysing efficiency [18, 23-25], assuming that lysis is directly proportional to the product of tumour cell (RAJI-19 in our case) and CAR-T cell concentration thereby ignoring saturation effects. However, due to the limited capacity of CAR-T cells, their lysing efficiency fundamentally saturates at a concentration higher than that required for optimal lysis. Specifically, for a fixed tumour cell concentration, CAR-T cell lysing efficiency increases and gradually converges to its maximum value with increasing CAR-T cell concentration (shown in Fig. 1E). We conjecture that saturated lysing efficiency triggers bistable RAJI-10 cell kinetics. This implies that within a given CAR-T cell concentration interval, RAJI-19 cell kinetics with low tumour burden are inhibited, but those with high tumour burden persist (Fig. 1F). This model prediction offers a compelling explanation for the clinically observed phenomenon that patients with higher tumour burdens at the start of treatment are less likely to both attain and maintain a deep response compared to those with lower tumour burden in B-ALL [1, 5, 11].

## Results

### CAR-T cell lysing efficiency increases and saturates with increasing concentrations of CAR-T cells and RAJI-19 cells

We quantified CAR-T cell lysing efficiency using a flow cytometry-based killing (FBK) assay for Raji-19 cells tested against VIII-CAR T cells or UT-CAR cells. This assay allowed the estimation of RAJI-19 cell and viable remaining RAJI-19 cell concentration before and over 72-hour incubation (see *Methods*) [28]. RAJI-19 cells were cocultured with UT-CAR and VIII-CAR T cells, manufactured from both healthy donors and B-ALL patients, at various effector-to target (T) cell ratio (E:T ratios: 1:1, 1:2, 1:4 and 1:8, shown in Fig.2) [28].

To identify the best-fit model for describing CAR-T cell lysing efficiency, we first used FBK assay data of RAJI-19 cells tested against UT-CAR cells from healthy donor at end-of-manufacture as an example (Section 1.1, *Model Selection*, Supplementary Material). To establish a quantitative relationship between RAJI-19 and CAR-T cell concentrations, we developed four distinct models:

- Unsaturated lysing model (System 1, *Methods*),
- Semi-saturated lysing model with increase of RAJI-19 cell concentration (System 2, *Methods*),
- Semi-saturated lysing model with increase of CAR T-cell concentration (System 3, *Methods*),
- Saturated lysing model with increase of RAJI-19 cell and CAR-T cell concentration (System 4, *Methods*).

These models were designed to describe the rates of change for both Raji-19 and CAR-T cell concentrations. Critically, the model allowing saturation with increasing Raji-19 and CAR-T cell concentrations (System 4, Methods, blue curve in Fig. 2B) consistently fit the FBK estimates (Fig. 2A and 2B). In contrast, unsaturated models (System 1, green curve in Fig. 2B) and semi-saturated models (System 2 and System 3, purple and light blue curve in Fig. 2B) failed to accurately capture FBK assay data (Table S1-S4, Model Selection, Supplementary Material). Furthermore, the saturated lysing model (System 4) consistently fit the FBK assay data across all experimental conditions, including UT-CAR and VIII-CAR T cells, at both end-of-manufacture and post-rechallenge states, and for cells derived from both healthy donors and B-ALL patients (Fig. 2C-H).

**Figure 2.**
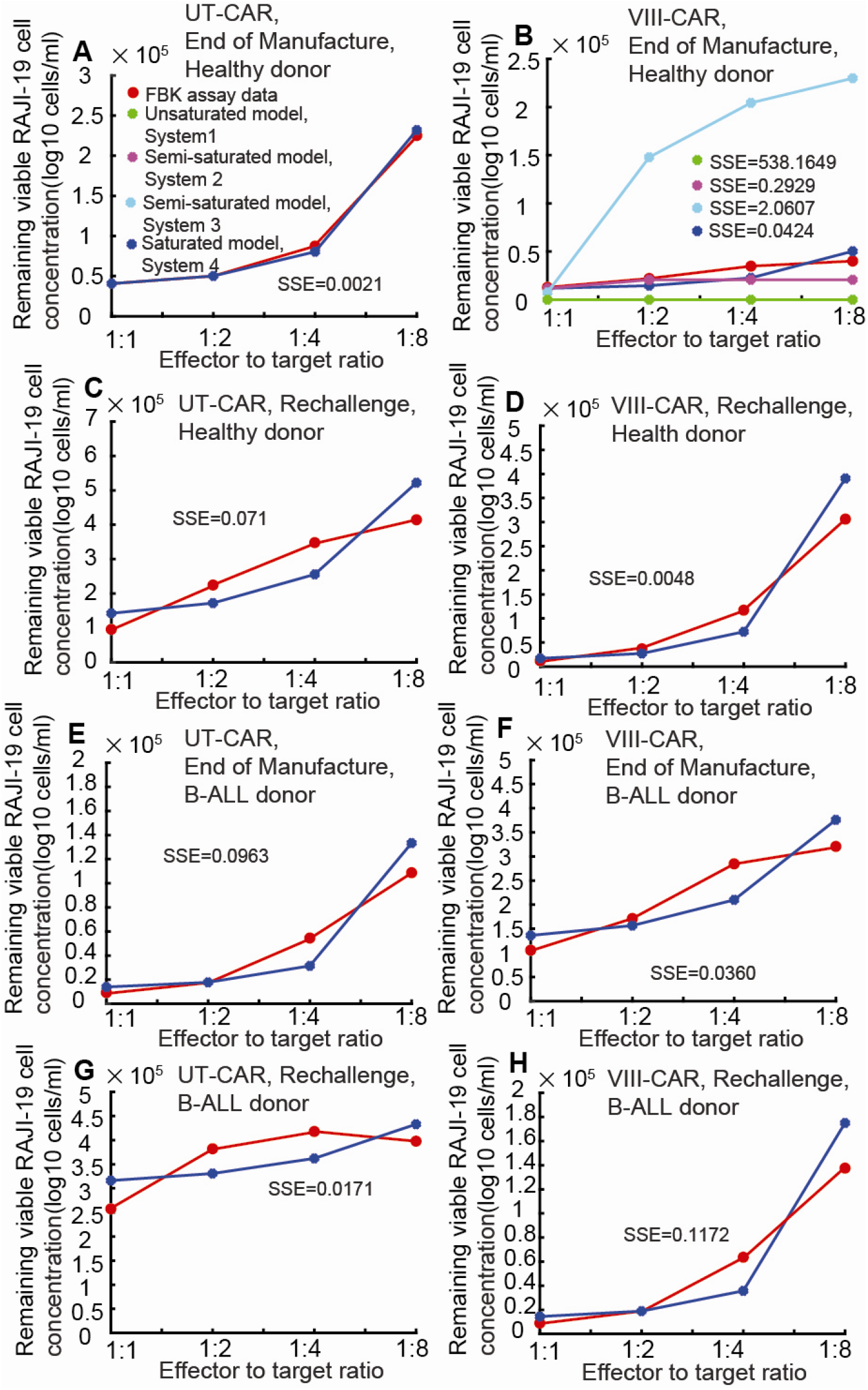
Quantitative relationship between RAJI-19 cell and CAR-T cell concentration with different E:T ratio established through FBK assay. Flow cytometry-based killing (FBK) assay data show that the remaining viable Raji-19 cell concentration generally decreases with an increasing effector-to-target (E:T) ratio (1:1, 1:2, 1:4, and 1:8) after 72 hours of incubation. **(A-D)** Data from CAR-T cells manufactured from healthy donors: **(A)** FBK assay of Raji-19 cells tested against UT-CAR T cells at the end-of-manufacture condition. **(B)** FBK assay of Raji-19 cells tested against VIII-CAR T cells at the end-of-manufacture condition. **(C)** FBK assay of Raji-19 cells tested against UT-CAR T cells post-rechallenge. **(D)** FBK assay of Raji-19 cells tested against VIII-CAR T cells post-rechallenge. **(E-H)** Data from CAR-T cells manufactured from B-ALL patients: **(E)** FBK assay of Raji-19 cells tested against UT-CAR T cells at the end-of-manufacture condition. **(F)** FBK assay of Raji-19 cells tested against VIII-CAR T cells at the end-of-manufacture condition. **(G)** FBK assay of Raji-19 cells tested against UT-CAR T cells post-rechallenge. **(H)** FBK assay of Raji-19 cells tested against VIII-CAR T cells post-rechallenge. Legend for panels **(A-H):** Red curves and circles represent the FBK assay data. Green curves and circles represent predictions provided by the unsaturated model (System 1). Purple curves and circles represent predictions provided by the semi-saturated model (System 2). Light blue curves and circles represent predictions provided by the semi-saturated model (System 3). Blue curves and circles represent predictions provided by the saturated model (System 4).

To understand CAR-T cell lysing efficiency across diverse experimental conditions (i.e., with and without AKT inhibitor VIII, at end-of-manufacture or post-rechallenge, and T cells derived from healthy donors or B-ALL patients), we compared three lysing parameters induced by different combinations of RAJI-19 cell. Generally, we observed that the lysing rate *κ* for VIII-CAR T cells was higher than that for UT-CAR T cells across most conditions.

For CAR-T cells derived from healthy donors, we observed:

- At the end-of-manufacture condition, lysing rate of UT-CAR T cell primarily ranged from 0 to 5 (Table. S4 in Supplementary Material), while for VIII-CAR T cells, it primarily ranged from 5 to 10 (Table. S5 in Supplementary Material). This finding suggests improved cytotoxicity for VIII-CAR T cells, which appears to contradict the conclusion of “comparable killing” noted in a previous study (Fig. 2B in Reference [28]).
- At the post-rechallenge condition, lysing rate of UT-CAR T cell primarily ranged from 4 to 8 (Table. S6, *Model Selection*, Supplementary Material), whereas for VIII-CAR T cells, it significantly increased, ranging from 20 to 30 (Fig. 3B, Table. S7, *Model Selection*, Supplementary Material). This latter finding is consistent with the observation that VIII-CAR displayed significantly improved cytotoxicity at all E:T ratios in the original biological study (Fig. 2B in Reference [28]).

**Figure 3.**
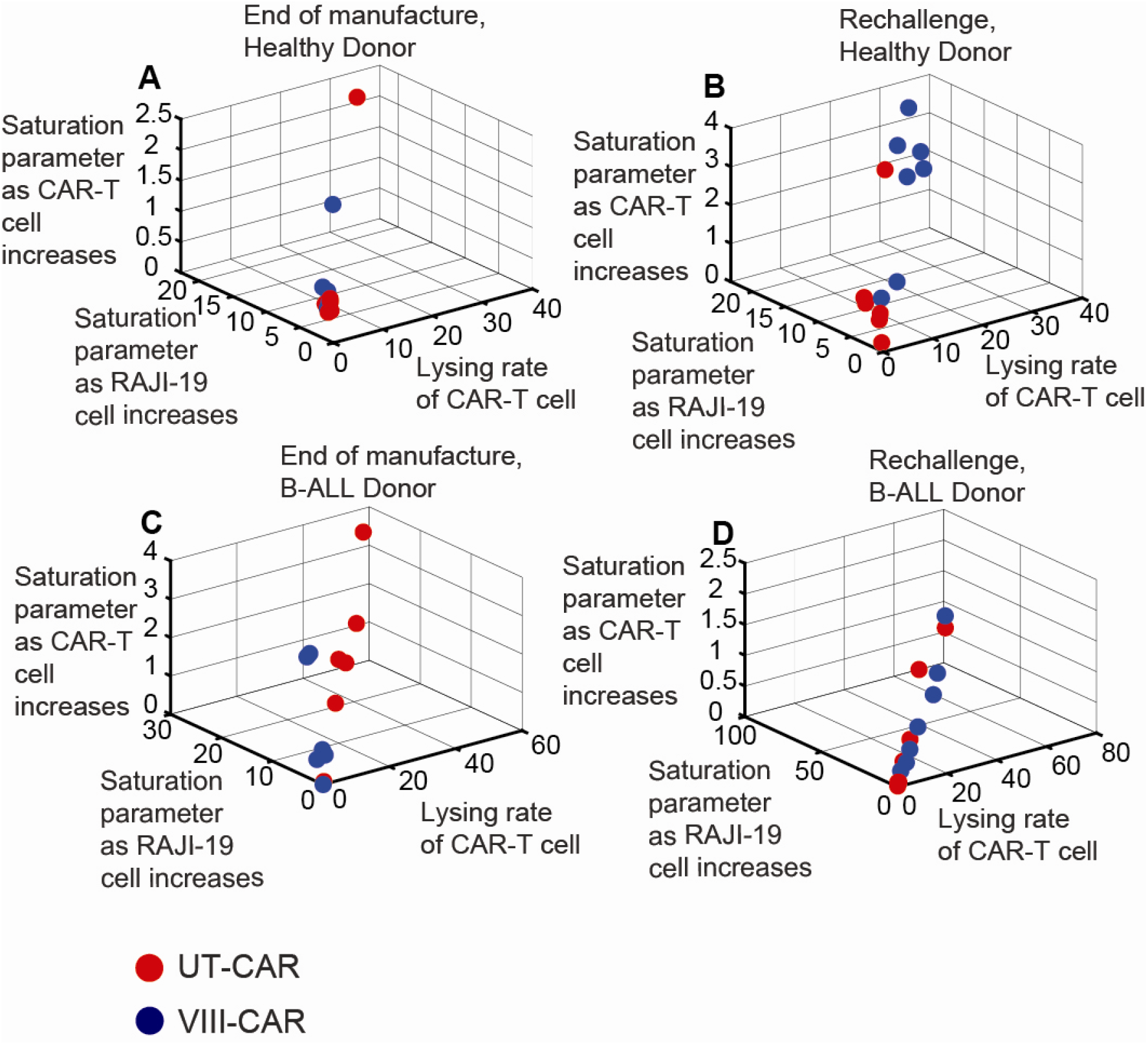
Distribution of killing parameters influenced by Raji-19 cells and CAR-T cells. This figure illustrates the estimated distributions for three key killing parameters derived from model fits for diverse experimental conditions (i.e., with and without AKT inhibitor VIII, at end-of-manufacture or post-rechallenge, and T cells derived from healthy donors or B-ALL patients) : **(A)** Parameters for Raji-19 cells tested against UT-CAR and VIII-CAR T cells, generated from healthy donors at the end-of-manufacture condition. **(B)** Parameters for Raji-19 cells tested against UT-CAR and VIII-CAR T cells, generated from healthy donors in the post-rechallenge condition. **(C)** Parameters for Raji-19 cells tested against UT-CAR and VIII-CAR T cells, generated from B-ALL donors at the end-of-manufacture condition. **(D)** Parameters for Raji-19 cells tested against UT-CAR and VIII-CAR T cells, generated from B-ALL donors in the post-rechallenge condition. In all panels, red circles represent data associated with UT-CAR T cells, while blue circles represent data associated with VIII-CAR T cells. The x-axis represents lysing of CAR-T cells. The y-axis represents saturation parameter as RAJI-19 cell concentration increases. The z-axis represents saturation parameter as CAR-T cell concentration increases.

For CAR-T cells derived from B-ALL patients: we observed:

- At the end-of-manufacture condition, lysing rate of VIII-CAR T cell primarily ranges from 0 to 10, while for UT-CAR T cell, it ranged from 20 to 40. This indicates that UT-CAR T cells exhibited better cytotoxicity in this specific scenario, a result that contradicts biological observations from Reference [28] and warrants further investigation.
- At the post-rechallenge condition, lysing rate for UT-CAR T cells primarily ranged from 0 to 10, and for VIII-CAR T cells, it ranged from 6 to 33. Therefore, for both end-of-manufacture and post-rechallenge conditions, VIII-CAR generally demonstrated significantly improved cytotoxicity for B-ALL patient-derived CAR-T cells, which is consistent with biological conclusions (Fig 5D in Reference [28]).

Expectedly, for T cells manufactured from both B-ALL patients and healthy donors, non-transduced T cells exhibited a low lysing rate (Table S8, Model Selection, Supplementary Material). This suggests that non-transduced T cells may not effectively inhibit Raji-19 cell replication, as anticipated.

### Transduced CAR-T Cells Lead to Bistable RAJI-19 Cell Kinetics, While Non-Transduced Cells Lead to Monostable Kinetics

In the preceding section, we observed distinct differences in CAR-T cell lysing efficiency among VIII-CAR T cells, UT-CAR T cells, and non-transduced cells. Utilizing bifurcation analysis and numerical simulations, we found that both VIII-CAR T cells and UT-CAR T cells led to bistable RAJI-19 cell kinetics, regardless of whether they were derived from healthy or B-ALL donors. In contrast, non-transduced T cells consistently resulted in monostable RAJI-19 cell kinetics. Notably, VIII-CAR T cells exhibited a smaller critical concentration required for RAJI-19 cell inhibition compared to UT-CAR T cells; this indicates that VIII-CAR T cells provide a superior therapy outcome. Bistability signifies that for a given CAR-T cell concentration, RAJI-19 cell kinetics with high tumour burdens persist, while those with low tumour burdens are suppressed (Fig. 1F). Conversely, monostability implies that the persistence or inhibition of RAJI-19 cell kinetics is solely determined by the magnitude of the CAR-T cell concentration (Fig. 6B).

To inform our simulations, we established a physiologically relevant range of CAR-T cell dosages. A typical adult has an approximate blood volume of 5 liters (5000 mL) and a body weight of 80 kg. Previous studies have proposed varying CAR-T cell dosages:

- **Brentjens et al**. [12] suggested a range of 1×10^9^ to 11.1×10^9^ cells, which translates to 2×10^5^ to 22.2×10^5^ cells/mL.
- **Gang Heng et al**. [38] proposed dosages of 12.3×10^6^ to 41.67×10^6^ cells/kg, equivalent to 1.96×10^5^ to 6.672×10^5^ cells/mL (assuming an 80 kg adult).
- For paediatric patients, **Heather Stefanski et al**. [8] recommended a single dose of tisagenlecleucel (a CD19-targeting CAR-T cell) 0.1 to 2.5 × 10^8^ CAR-T cells/kg for patients >50 kg.

Assuming blood volumes ranging from 3250 mL (female adolescent) to 3500 mL (male adolescent), and an estimated CAR-T cell infusion number from 1.5 ×10^5^ cells/mL to 35×10^5^ cells/mL, we selected a CAR-T cell dosage range of 3×10^5^ cells/mL to 20×10^5^ cells/mL for model.

We first utilized FBK assay data from post-rechallenge VIII-CAR T cells, manufactured from healthy donors, as an illustrative example. The specific lysing parameters used in these simulations were sourced from column three of Table S7 (Section 1.1, *Supplementary Material*).

In simulations of Raji-19 cell growth kinetics with saturated killing efficiency (System 4), a critical threshold (C*=9.78×10^5^ cells/mL) divided the CAR-T cell concentration interval into two distinct dynamic regions, as depicted in the bifurcation diagram (Fig. 4D). At different VIII-CAR T cell concentration intervals, RAJI-19 cell kinetics exhibited different dynamics:

**Figure 4.**
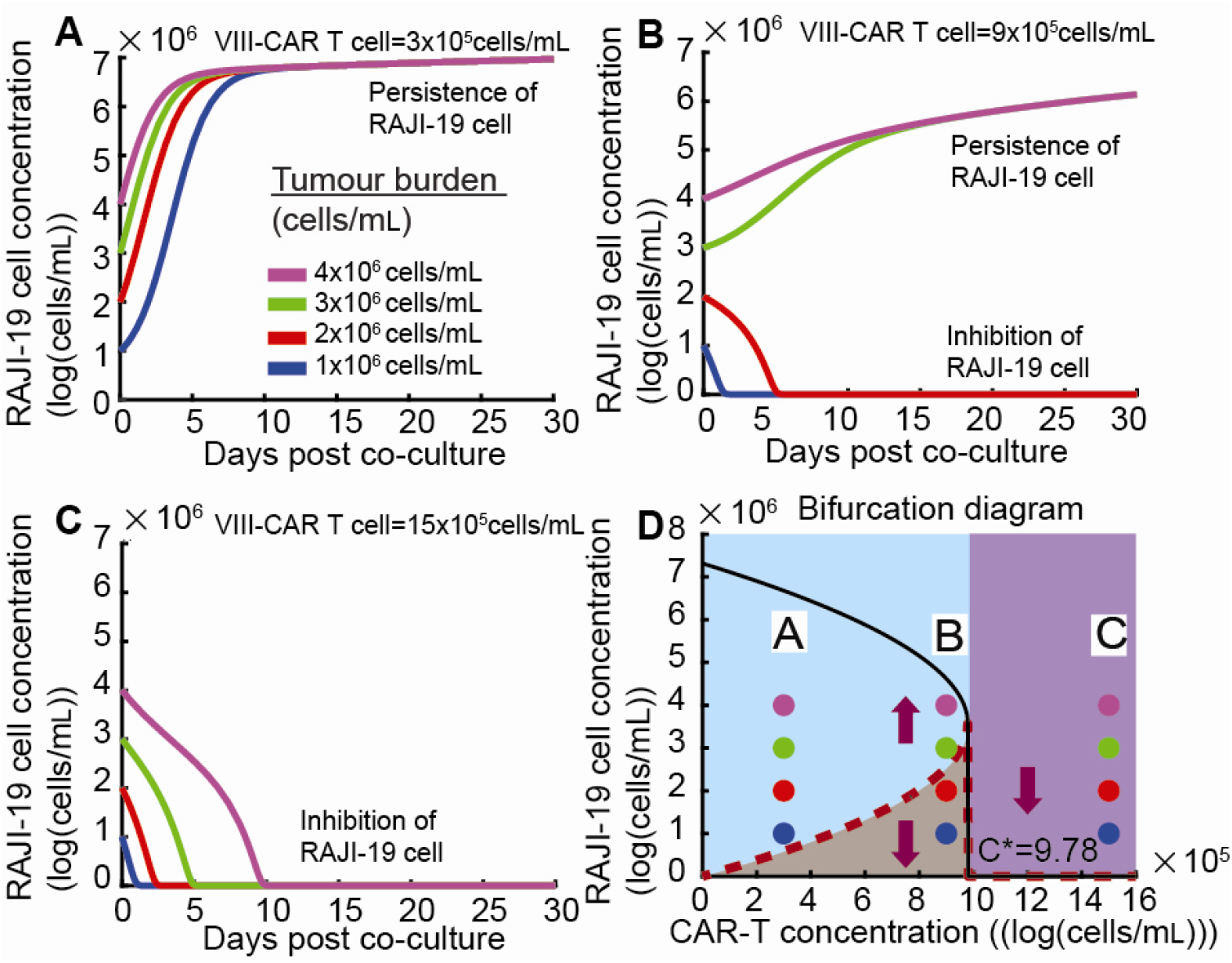
Simulated Kinetics of RAJI-19 Cells under Varying Tumour Burdens and VIII-CAR T Cell Concentrations. This figure visualizes the simulated Raji-19 cell kinetics demonstrating bistable behaviours based on different tumour burdens and post-rechallenge VIII-CAR T cell concentrations (manufactured from health donors). **(A, B)** These panels illustrate RAJI-19 cell growth kinetics when the VIII-CAR T cell concentration is within the range of 0 and C*. In this interval, RAJI-19 cell growth remains unaffected if the tumour burden is above the dashed red curve (threshold for inhibition), but it is inhibited if the initial tumour burden falls below this curve. **(C)** This panel shows that RAJI-19 cell kinetics, regardless of tumour burdens, are completely eradicated when the VIII-CAR T cell concentration exceeds C*. **(D)** A bifurcation diagram depicting the steady-state RAJI-19 concentration as a function of VIII-CAR T cell concentration. The tumour burden threshold (red dashed curve) for inhibition increases with increasing VIII-CAR T cell concentration. The maximal capacity of RAJI-19 concentration (black solid curve) decreases as VIII-CAR T cell concentration increases. Purple arrows in the bifurcation diagram **(D)** indicate the dynamic behaviour for any given tumour burden. Blue, red, green, and purple lines and circles in panels (A-D) represent the simulated Raji-19 cell kinetics for initial tumour burdens of 1×10^6^ cells/mL, 2×10^6^ cells/mL, 3×10^6^ cells/mL and 4×10^6^ cells/mL in (A–D), respectively. System 5 was used for these numerical simulations, utilizing the following parameters: *κ* = 29.42, *γ* = 16.66, *η* = 2.87 and *φ* = 0.3 × 10^−10^.

**Figure 5.**
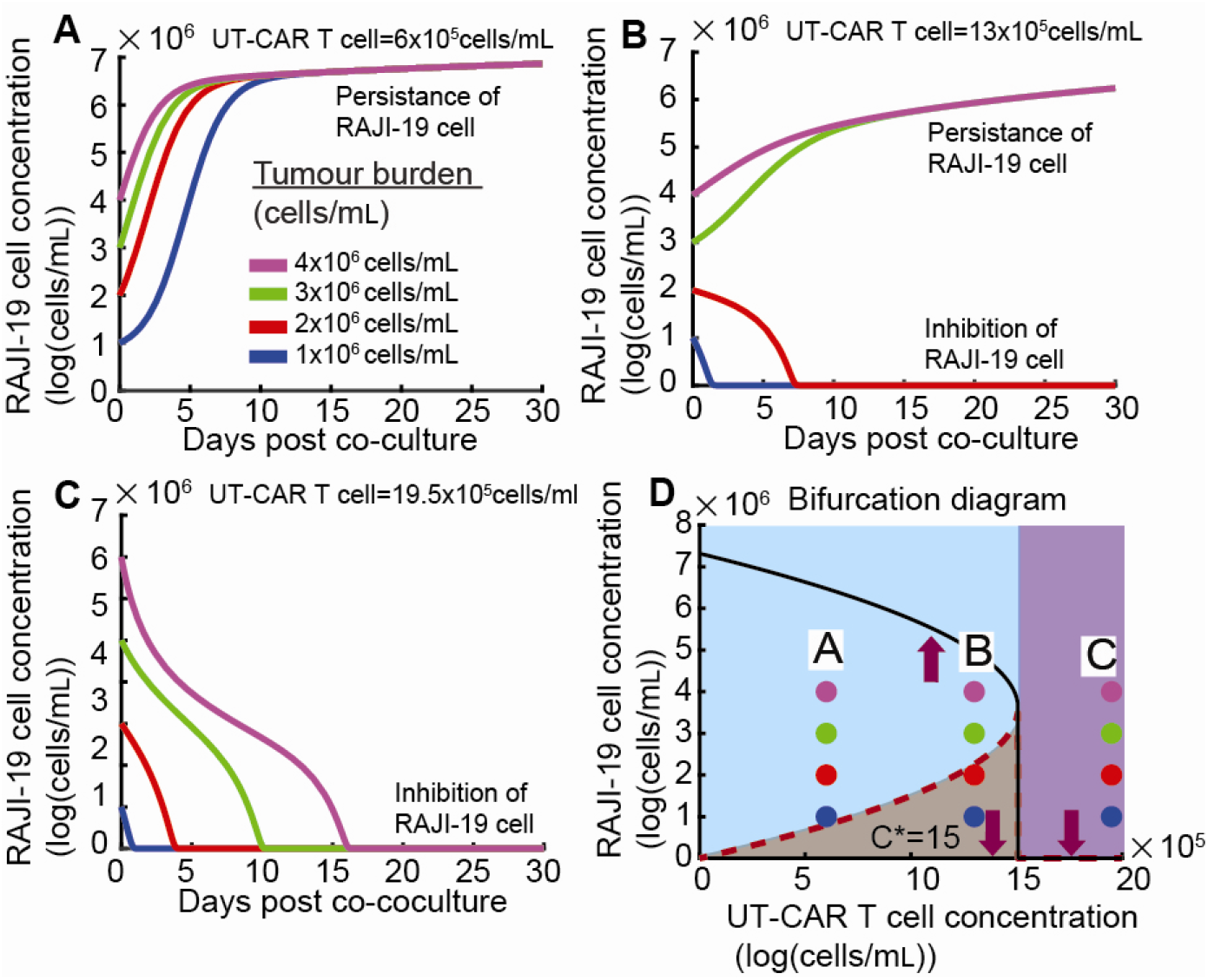
Simulated Kinetics of RAJI-19 Cells Under Varying Tumour Burdens and UT-CAR T Cell Concentrations. This figure visualizes the simulated Raji-19 cell kinetics, demonstrating the impact of different tumour burdens and post-rechallenge UT-CAR T cell concentrations (from healthy donors). **(A, B)** These panels illustrate RAJI-19 cell growth kinetics when the UT-CAR T cell concentration is within the range of 0 and C*. In this interval, Raji-19 cell growth isn’t affected if the tumour burden is above the dashed red curve (threshold for eradication), but it’s eradicated if the tumour burden falls below this curve. **(C)** This panel shows that Raji-19 cell growth kinetics, regardless of tumour burden, are inhibited when the UT-CAR T cell concentration exceeds C*. **(D)** A bifurcation diagram depicting the steady-state RAJI-19 concentration as a function of UT-CAR T cell concentration. The tumour burden threshold (red dashed curve) for inhibition increases with increasing UT-CAR T cell concentration. The maximal capacity of RAJI-19 concentration (black solid curve) decreases as UT-CAR T cell concentration increases. Purple arrows in the bifurcation diagram indicate the dynamic behaviour for any given tumour burden. Blue, red, green, and purple lines and circles in panels (A-D) represent the simulated Raji-19 cell kinetics for tumour burdens of 1×10^6^ cells/mL, 2×10^6^ cells/mL, 3×10^6^ cells/mL and 4×10^6^ cells/mL in (A–D), respectively. System 5 was used for these numerical simulations, utilizing the following parameters: *κ* = 4.96, *γ* = 4.39, *η* = 0.31 and *φ* = 0.3 × 10^-10^.

**Figure 6.**
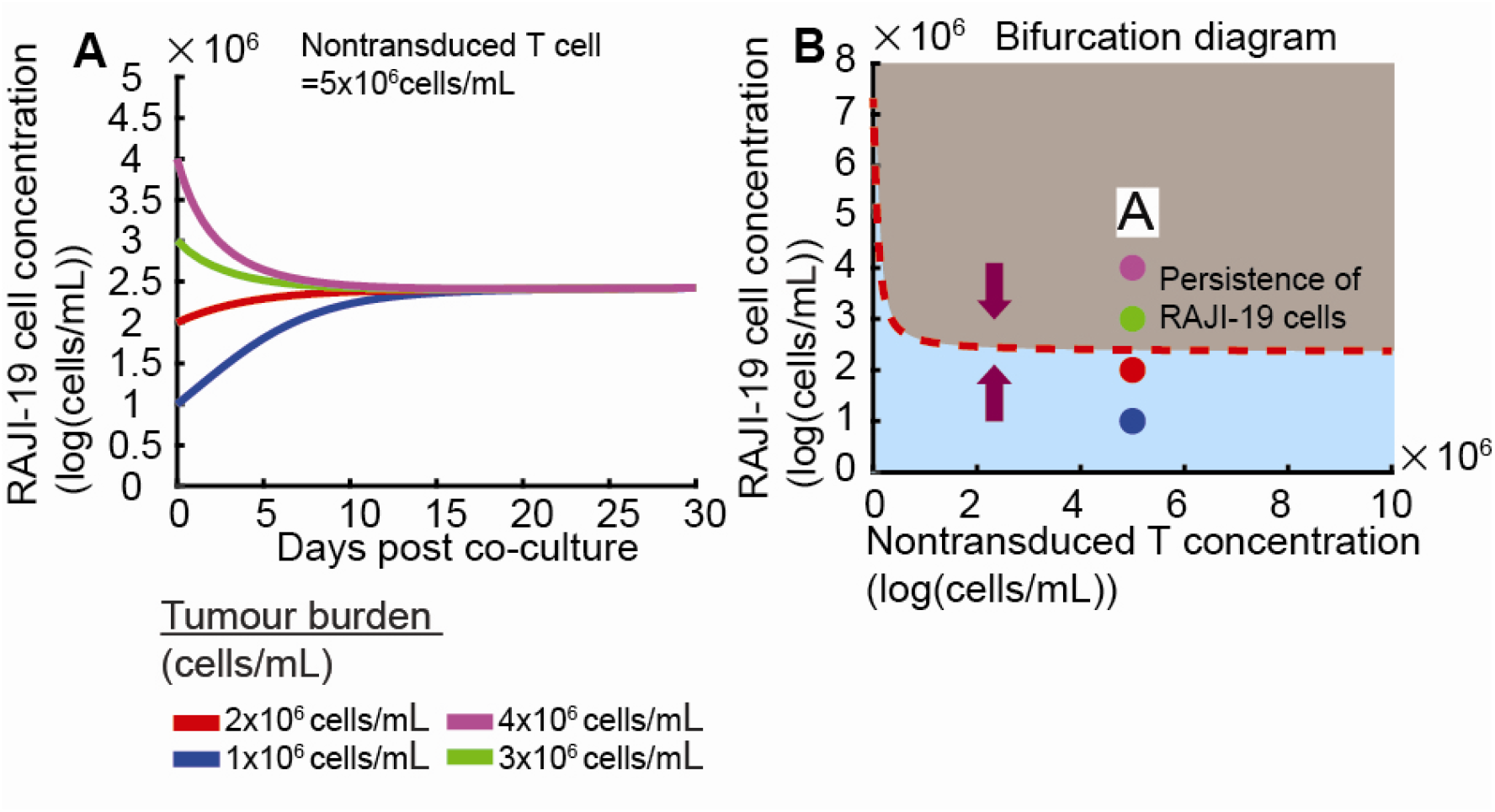
Simulated Kinetics of Raji-19 Cells Under Varying Tumour Burdens and Non-Transduced T Cell Concentration. This figure illustrates the simulated Raji-19 cell kinetics, demonstrating the impact of different combinations of tumour burdens and non-transduced T cell (NT) concentrations, which consistently result in monostable behaviours. **(A)** RAJI-19 cell kinetics consistently converge to a unique steady state, regardless of tumour burden. **(B)** A bifurcation diagram illustrating the RAJI-19 cell concentration as a function of non-transduced T cell concentration. The purple arrows in the diagram represent the dynamic behaviour for any given tumour burden, consistently pointing towards a single stable equilibrium. Blue, red, green, and purple lines and circles in panels (A-D) represent the simulated Raji-19 cell kinetics for tumour burdens of 1×10^6^ cells/mL, 2×10^6^ cells/mL, 3×10^6^ cells/mL and 4×10^6^ cells/mL in (A–B), respectively. System (5) was used to perform this numerical simulation. System 5 was used for these numerical simulations, utilizing the following parameters *κ* = 1.79, *γ* = 2.92, *η* = 0.051 and *φ* = 0.3 x 10^-10^.

- **Bistability**: Raji-19 cell kinetics displayed bistability in the CAR-T cell concentration interval between 0 and C*. In this region, Raji-19 cells with low tumour burdens were inhibited, while those with high tumor burdens persisted, even at the same VIII-CAR T cell concentration (Fig. 4B and 4D). The Raji-19 cell concentration threshold (above which the Raji-19 cell concentration remains unaffected) was observed to increase with rising VIII-CAR T cell concentration (red dashed curve in Fig. 4D). For instance, at a low VIII-CAR T cell concentration (C=3 ×10^5^ cells/mL), Raji-19 tumour burdens of 1×10^6^ cells/mL, 2×10^6^ cells /mL, 3×10^6^ cells /mL and 4×10^6^ cells /mL all persist (Fig. 4A and 4D). In contrast, at a higher VIII-CAR T cell concentration (9×10^5^ cells/mL), only high tumour burdens of 3×10^6^ cells /mL and 4×10^6^ cells /mL persists (Fig. 4B and 4D).
- **Monostability**: At VIII-CAR T cell concentrations higher than the threshold (C*=9.78×10^6^ cells/mL), Raji-19 cell growth was inhibited regardless of the initial tumour burden (Fig. 4C and 4D).

It’s also worth noting that VIII-CAR T cell concentration kinetics decreased after days post-infusion (Fig. S1).

The degree of CAR-T cell proliferation varies significantly among donors, with reported increases ranging from 2-3 fold in some individuals to over 20-fold in others during a seven-day culture period [39]. The example presented above utilized a low rate of proliferation and exhaustion of CAR T-cells when stimulated by cancer cells *φ* = 0.3 × 10 ^−10^ (*day* ^− 1^ *cell* ^− 1^) as provided by Reference [18]. We subsequently found that regardless of infusion cell concentration, RAJI-19 cell kinetics was inhibited with high rate of proliferation and exhaustion of CAR-T cell *φ* = 0.3 × 10 ^−7^ (*day* ^− 1^ *cell* ^− 1^). This inhibition occurred because the resulting CAR-T cell concentration remained consistently higher than the critical threshold (C*=9.78×10^6^ cells/mL) (shown in Fig. S3).

Similarly, we performed numerical simulations using FBK assay data from post-rechallenge UT-CAR T cells, manufactured from healthy donors. The lysing parameters for these simulations were obtained from column one of Table S6 (Section 1.2, *Supplementary Material*). Our simulations revealed that UT-CAR T cells also induce bistable Raji-19 cell kinetics (Fig. 5). However, the critical concentration threshold (C*=15×10^5^ cells/mL) induced by UT-CAR T cells is considerably higher than the threshold observed with VIII-CAR T cells (Fig. 4). This notable difference suggests that VIII-CAR T cells may offer a superior therapy outcome due to their ability to achieve tumour inhibition at lower critical concentrations. Furthermore, similar to VIII-CAR T cells, the UT-CAR T cell concentration kinetics also showed a decrease after days post-infusion (Fig. S2).

Finally, our analysis revealed that non-transduced T cells (NT) consistently lead to monostable RAJI-19 cell kinetics (Fig. 6). This signified that even significantly high concentrations of non-transduced T cells are unable to inhibit RAJI-19 cell growth. This outcome is consistent with previous observations where non-transduced T cells typically exhibit less than 5–10% cytotoxicity and are consequently used as a negative control in studies like Reference [28]. In contrast, transduced CAR-T cells (e.g., VIII-CAR and UT-CAR) showed approximately 80% cytotoxicity in Reference [28]. This stark difference in lytic capability explains the observed phenomenon: non-transduced T cells induced monostability, whereas transduced CAR-T cells induced bistability. Therefore, despite any potential proliferation, non-transduced T cells cannot inhibit RAJI-19 cell growth. The lysing parameters for this simulation were taken from row three of Table S8 (Section 1.2, *Supplementary Material*).

To summarize, transduced CAR-T cells consistently led to bistable RAJI-19 cell kinetics, a phenomenon where tumour eradication depends on tumour burden and CAR-T cell concentration. Notably, VIII-CAR T cells provided a smaller threshold value for R cell eradication, suggesting better treatment outcomes compared to UT-CAR T cells (Table 1). The CAR-T cell state (end-of-manufacture or post-rechallenge) did not alter whether the kinetics were bistable or monostable. However, it did significantly influence the specific threshold values required for effective treatment outcomes (fifth column in Table 1). In contrast, non-transduced T cells consistently resulted in monostable RAJI-19 cell kinetics, regardless of whether they were from the end-of-manufacture or rechallenge conditions. This indicates that non-transduced T cells are unable to provide effective therapeutic outcomes (Table 1).

**Table 1.**
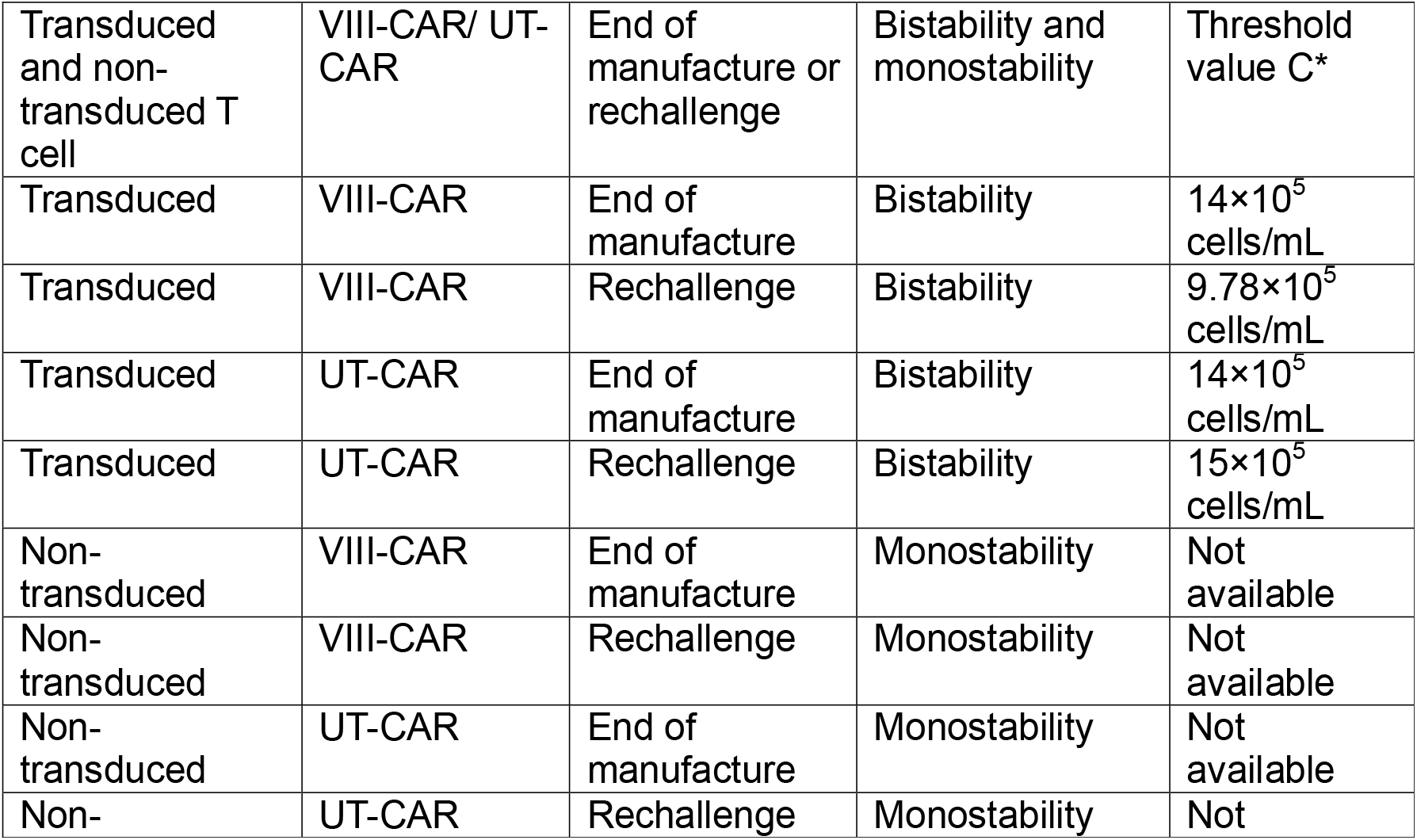
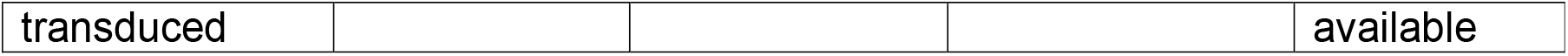
Bistability and monostability determined by non-transduced/transduced T CAR cell.

### CAR-T Cell Proliferation and Dosing Regimens Determine Raji-19 Cell Kinetics

In the previous section, we established that CAR-T cell-induced bistable RAJI-19 cell kinetics are responsible for observed variations in therapy outcomes. Specifically, with a single-dose infusion, RAJI-19 cell kinetics with high tumour burdens persist, while those with low tumour burdens are inhibited. Administering split-dose infusions of CAR-T cells over 2-3 days, instead of a single-dose infusion, has been shown to reduce the toxicity of CAR-T cell therapy, particularly cytokine release syndrome (CRS) and neurotoxicity [40]. Here, we investigate how different split-dose infusion scenarios influence CAR-T cell therapy outcomes.

Based on existing literature, we considered different split-dose infusion scenarios:

- **Three-dose split infusion:** Noelle V Frey al et [41], David L. Porter al et [4] and Michael Kalos al et [42] proposed a regimen where the first infusion contains 10% of the total cells, the second 30%, and the third 60%.
- **Two-dose split infusion:** Mark B. Geyer al et [43] and Craig S. Sauter al et [44] proposed a regimen where the first infusion contains 33% of the cells and the second contains 67%.

Building on these, we designed and analyzed four distinct treatment scenarios (Fig. 7A):

**Figure 7.**
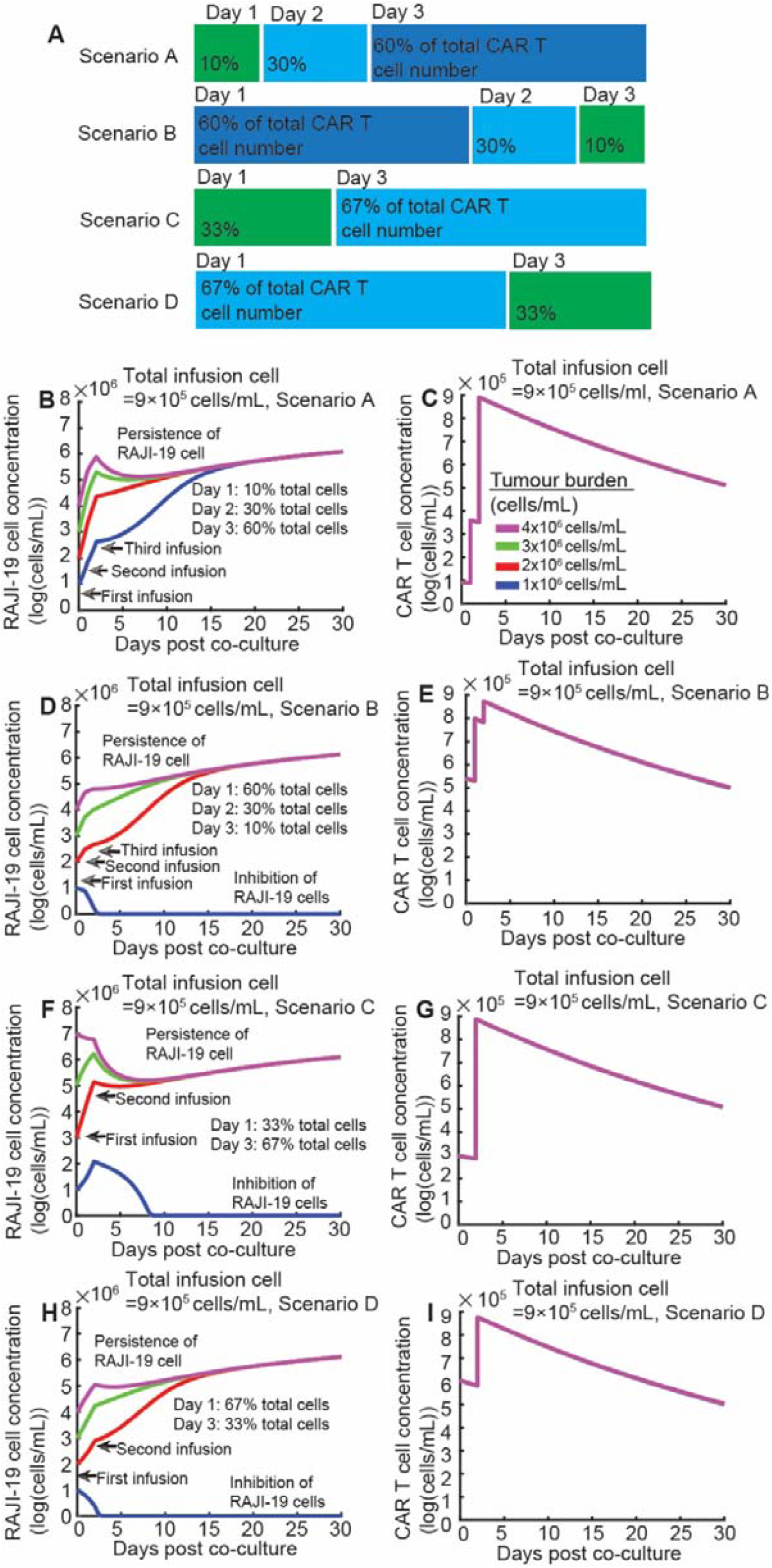
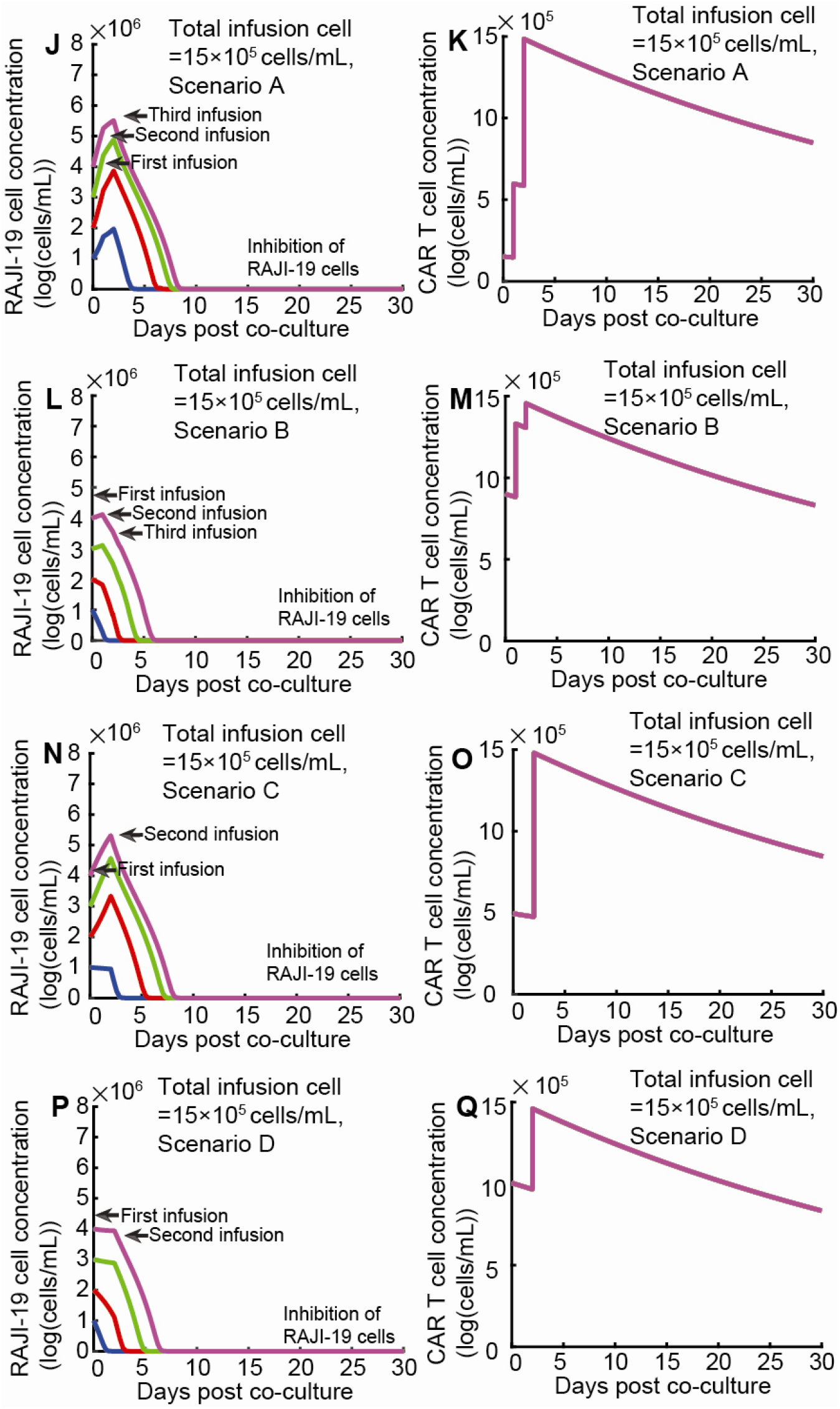
Simulated kinetics of Raji-19 cells with a low CAR-T cell proliferation rate, exploring different combinations of tumour burden and CAR-T cell infusion strategies. This figure presents simulated Raji-19 cell kinetics under various split-dose CAR-T cell infusion scenario. **(A) Schematic diagram of split infusion scenarios: Scenario A:** 10% of total cells infused on Day 1, 30% on Day 2, and 60% on Day 3. **Scenario B:** 60% of total cells infused on Day 1, 30% on Day 2, and 10% on Day 3. **Scenario C:** 33% of total cells infused on Day 1 and 67% on Day 3. **Scenario D:** 67% of total cells infused on Day 1 and 33% on Day 3. **(B-I) Simulations with a total infusion cell concentration of 9×10**^**6**^ **cells/mL: (B) Raji-19 cell kinetics** and **(C) CAR-T cell kinetics** under **Scenario A. (D) Raji-19 cell kinetics** and **(E) CAR-T cell kinetics** under **Scenario B. (F) Raji-19 cell kinetics** and **(G) CAR-T cell kinetics** under **Scenario C. (H) Raji-19 cell kinetics** and **(I) CAR-T cell kinetics** under **Scenario D. (J-Q) Simulations with a total infusion cell concentration of 15×10**^**6**^ **cells/mL: (J) Raji-19 cell kinetics** and **(K) CAR-T cell kinetics** under **Scenario A. (L) Raji-19 cell kinetics** and **(M) CAR-T cell kinetics** under **Scenario B. (N) Raji-19 cell kinetics** and **(O) CAR-T cell kinetics** under **Scenario C. (P) Raji-19 cell kinetics** and **(Q) CAR-T cell kinetics** under **Scenario D**. In panels (B), (D), (F), (H), (J), (L), (N), and (P), **purple, green, red, and blue lines and circles** represent Raji-19 cell kinetics with tumour burdens of 1×10^6^ cells/mL, 2×10^6^ cells/mL, 3×10^6^ cells/mL, and 4×10^6^ cells/mL, respectively. In panels (C), (E), (G), (I), (K), (M), (O), and (Q), the **purple, green, red, and blue curves overlap** because the initial CAR-T cell concentrations for the different tumour burdens are the same within each specific scenario and total infusion concentration. System (5) was used to perform this numerical simulation. System 5 was used for these numerical simulations, utilizing the following parameters: *κ* = 29.42, *γ* = 16.66, *η* = 2.87 and *φ* = 0.3 x 10^-10^.

- **Scenario A:** Three-dose split infusion with 10%, 30%, and 60% of total cells in the first, second, and third infusions, respectively.
- **Scenario B:** Three-dose split infusion with 60%, 30%, and 10% of total cells in the first, second, and third infusions, respectively (reverse order).
- **Scenario C:** Two-dose split infusion with 33% and 67% of total cells in the first and second infusions, respectively.
- **Scenario D:** Two-dose split infusion with 67% and 33% of total cells in the first and second infusions, respectively (reverse order).

Given the decisive role of rate of proliferation and exhaustion of CAR-T cell play a decisive role in CAR-T therapy outcomes, we first analysed its effect. The value for rate of proliferation and exhaustion *φ* = 0.3 × 10 ^−10^ (*day* ^− 1^ *cell* ^− 1^) was adopted from Reference [18].

Unexpectedly, with a low infusion concentration (9×10^5^ cells/mL), two- or three-dose split infusion did not provide as favourable therapy outcomes as single-dose infusion, regardless of scenarios (shown in Fig. 7B-7I).

- **Scenario A** failed to inhibit RAJI-19 cells, irrespective of tumour burdens (shown in Fig. 7B)
- **Scenario B** could only inhibit low tumour burdens (e.g., 10^5^ cells/mL, shown in Fig. 7D). This limited efficacy was attributed to very low CAR-T cell concentrations during the first five days post-infusion.
- **Scenario C** failed to inhibit RAJI-19 cells, irrespective of tumour burdens (shown in Fig. 7F).
- **Scenario D** could only inhibit low tumour burden (e.g., 10^5^ cells/mL, shown in Fig. 7H).

In contrast, a single-dose infusion with a CAR-T cell concentration of 9×10^5^ cells/mL was capable of inhibiting tumour burden of 1×10^5^ cells/mL and 2×10^5^ cells/mL (shown in Fig. 4B and 4D). However, with a high total infusion concentration (15×10^5^ cells/mL), RAJI-19 kinetics is inhibited independent of four scenarios (shown in Fig.7J-7Q).

Furthermore, we found that the E:T ratio is a strong predictor for determining the inhibition or persistence of RAJI-19 cell, a finding that agrees with Reference [45].

- For single-dose infusion, RAJI-19 cell kinetics with low tumour burdens were inhibited, and their E:T ratio was greater than 1 (shown in Fig. S4A and S4B, blue and red curve).
- Conversely, for forward-order split-dose infusion, RAJI-19 cell kinetics persisted when their E:T ratio was less than 1 (shown in Fig. S4C and S4D).
- For reverse-order split-dose infusion, RAJI-19 cell kinetics with low tumour burdens were inhibited, when their E:T ratio is greater than 1 (shown in Fig. 4E and 4F, blue curve).

Next, we examined the impact of a higher rate of proliferation and exhaustion of CAR-T cell *φ* = 0.3 × 10 ^−7^ (*day* ^− 1^ *cell* ^− 1^). Even with a low total infusion concentration (9×10^5^ cells/mL), RAJI-19 cell kinetics were inhibited across all four scenarios. This occurred because the CAR-T cells rapidly proliferated, and their concentration remained consistently high (shown in Fig. 8).

**Figure 8.**
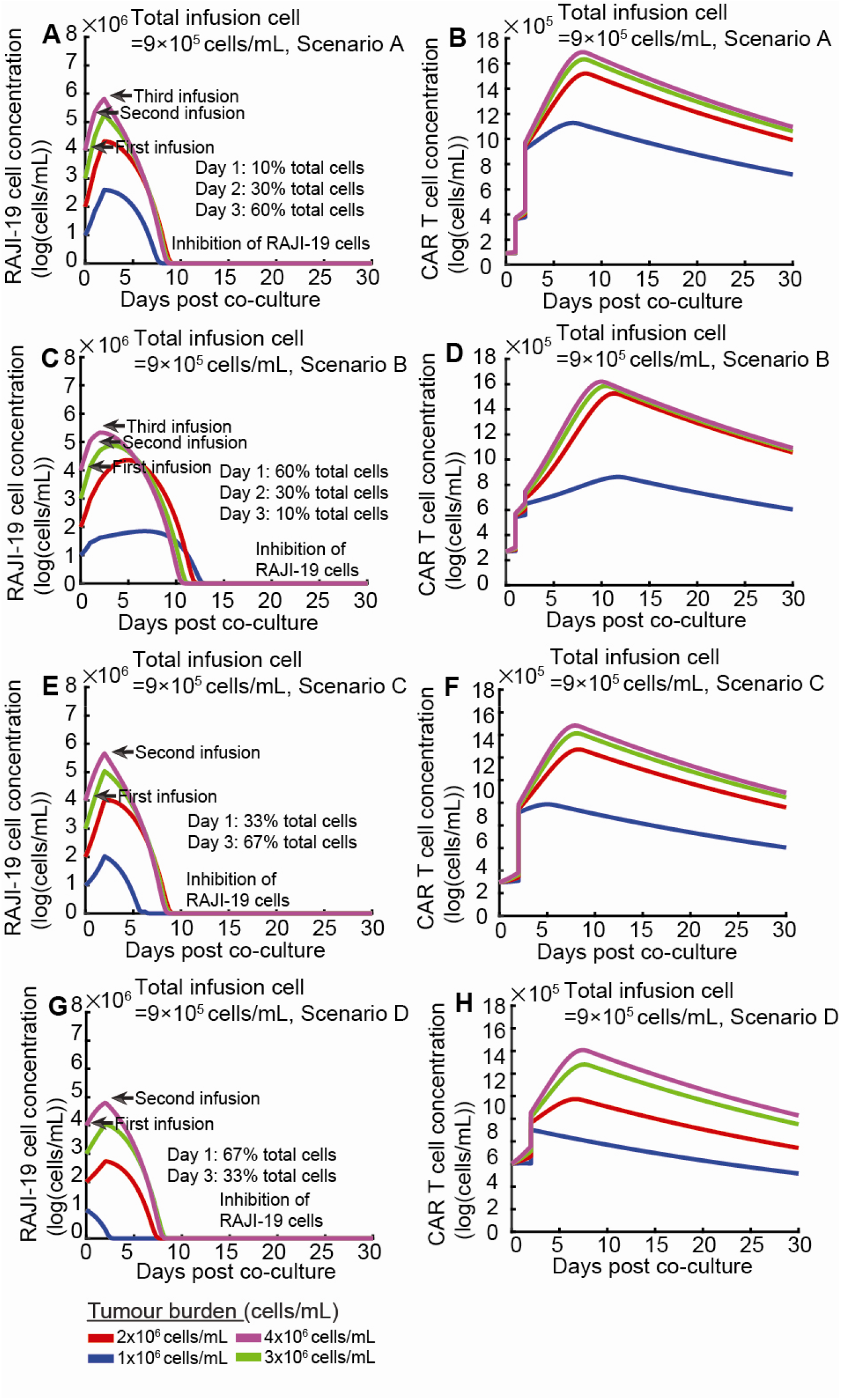
Simulated kinetics of Raji-19 cells with a high CAR-T cell proliferation rate, exploring different combinations of tumour burden and CAR-T cell infusion strategies. This figure presents simulated RAJI-19 cell kinetics under various split-dose CAR-T cell infusion strategies, specifically illustrating the impact of a high CAR-T cell proliferation rate. **(A) RAJI-19 cell kinetics** and **(B) CAR-T cell kinetics** under **Scenario A**: The three-dose split infusion strategy where 10% of cells are received on Day 1, 30% on Day 2, and 60% on Day 3. **(C) RAJI-19 cell kinetics** and **(D) CAR-T cell kinetics** under **Scenario B**: The three-dose split infusion strategy where 60% of cells are received on Day 1, 30% on Day 2, and 10% on Day 3. **(E) RAJI-19 cell kinetics** and **(F) CAR-T cell kinetics** under **Scenario C**: The two-dose split infusion strategy where 33% of cells are received on Day 1 and 67% on Day 3. **(G) RAJI-19 cell kinetics** and **(H) CAR-T cell kinetics** under **Scenario D**: The two-dose split infusion strategy where 67% of cells are received on Day 1 and 33% on Day 2. In panels (A), (C), (E), and (G), **purple, green, red, and blue lines and circles** represent Raji-19 cell kinetics for initial pre-infusion tumour burdens of 1×10^6^ cells/mL, 2×10^6^ cells/mL, 3×10^6^ cells/mL, and 4×10^6^ cells/mL, respectively. In panels (B), (D), (F), and (H), the **purple, green, red, and blue curves often overlap** because the initial CAR-T cell concentrations across different tumour burdens are the same within each specific scenario and total infusion concentration. System (5) was used to perform this numerical simulation. System (5) was used to perform this numerical simulation. Parameters *κ* = 29.42, *γ* = 16.66, *η* = 2.87 and *φ* = 0.3 x 10^-7^.

In summary, the rate of proliferation and exhaustion of CAR-T cells and the total infusion cell concentration directly determine treatment outcomes (Table 2). Specifically, a high rate of proliferation and exhaustion consistently leads to favourable treatment outcomes, irrespective of whether a split- or single-dose infusion is used. For a low rate of proliferation and exhaustion combined with a low total infusion concentration, a single-dose infusion provides better therapy outcomes. These conclusions are based on numerical simulations and warrant validation through further experimental studies.

**Table 2.**
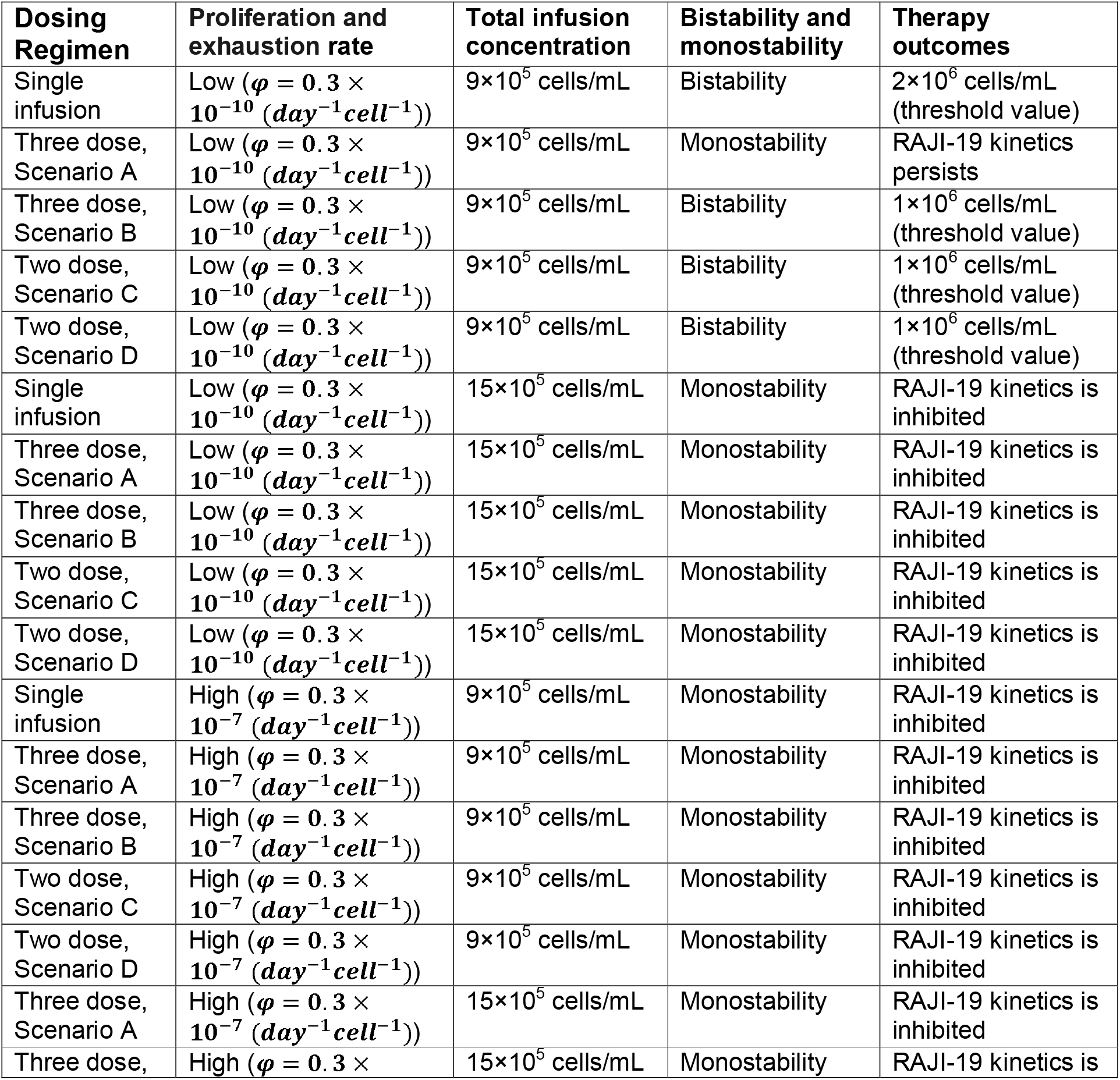
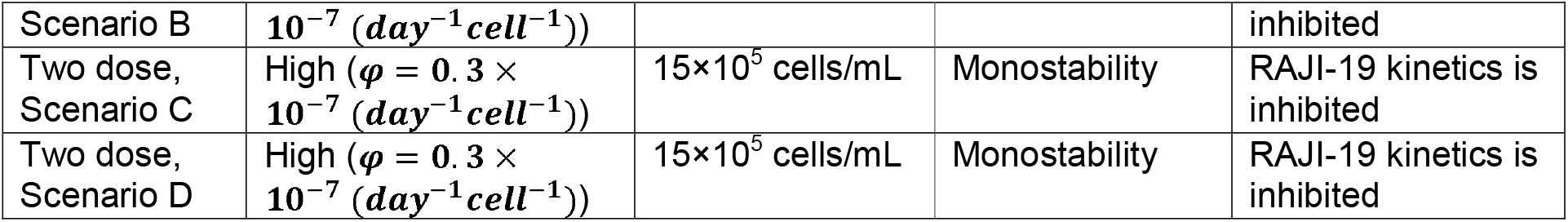
RAJI-19 cell Kinetics Determined by Infusion Dose, Dosing Regimen and Proliferation.

### Extrapolations to the *In Vivo* System

Our findings provide crucial insights into CAR-T cell therapy, offering direct extrapolations to the in vivo system.

- **CAR-T Cell Proliferation Predicts Outcomes**: Our findings suggest that CAR-T cell proliferation is the best predictor for the persistence or inhibition of RAJI-19 cell kinetics. When CAR-T cell proliferation is high, RAJI-19 cell kinetics are inhibited, regardless of the infusion dosage, tumour burden, or dosing regimens (single-dose or split-dose infusion). Conversely, when CAR-T cell proliferation is low, the infusion dosage, tumour burden, and dosing regimens jointly determine the persistence or inhibition of Raji-19 cell kinetics. This aligns with clinical observations that in vivo peak CAR-T cell expansion is the best predictor for short-term response[45].
- **CAR-T cell-induced bistable RAJI-19 kinetics and pretreatment tumour burden:** CAR-T cell-induced bistable RAJI-19 kinetics refers to the phenomenon where RAJI-19 cell kinetics with a high pre-infusion tumour burden are not cleared, while those with a low pre-infusion tumour burden are effectively controlled. This also matches the clinical observation that pretreatment disease burden was a useful predictor for remission duration and survival [45].
- **E:T Ratio Match Clinical Observations**: The E:T ratio is strongly associated with the persistence or inhibition of RAJI-19 cell kinetics. Specifically, regardless of whether a single-dose or split-dose infusion is used, an E:T ratio > 1 leads to the inhibition of RAJI-19 cell kinetics, whereas an E:T ratio < 1 induces the persistence of RAJI-19 cell kinetics. This is consistent with clinical observations showing that a higher ratio of peak CAR-T cell expansion to tumour burden significantly correlated with overall survival and was a better predictor of long-term survival [45].
- **Alignment with Clinical CAR-T Cell Concentrations and Dosing Insights**: Our predicted bistable CAR-T cell concentration interval aligns well with observed post-infusion CAR-T cell concentration intervals. Our predicted CAR-T cell concentration threshold corresponding to bistable RAJI-19 cell kinetics ranges from 9-14×10^5^ cells/mL. Fig. 1E in Reference [36] suggests that absolute CAR-T cell concentrations after proliferation range from 39.7 to 11,346 cells/μL (equivalent to 0.397-11.346× 10^5^ cells/mL), with a medium of 480.5 cells/μL (equivalent to 4.80× 10^5^ cells/mL). Similarly, Fig. 3C in Reference [29] suggests absolute CAR-T cell concentrations after proliferation range from 10^2^ to 10^4^ cells/μL (equivalent to 1-10× 10^5^ cells/mL), with a geometric mean of 1098.79 cells/μL (equivalent to 10.98× 10^5^ cells/mL).

We found that a single-dose infusion can perform better than a split-dose infusion even when the total number of infused cells is the same. The cell number in the first infusion appears to be a critical determinant of CAR-T cell therapy outcomes. This finding warrants further experimental validation.

## Discussion

Our analysis of flow cytometry-based killing (FBK) assay data demonstrated that the lysing efficiency of CAR-T cells (both VIII-CAR and UT-CAR) increases and then saturates with increasing concentrations of both RAJI-19 target cells and CAR-T cells. This saturation is key to the bistable RAJI-19 cell kinetics induced by both VIII-CAR and UT-CAR T cells. In contrast, non-transduced T cells induce monostable RAJI-19 cell kinetics, indicating their ineffective lysis of RAJI-19 cells and thus inefficient treatment outcomes. Importantly, VIII-CAR T cells exhibited a smaller critical CAR-T cell concentration threshold for favourable therapy outcomes compared with UT-CAR T cells. Furthermore, a high CAR-T cell proliferation rate, when stimulated by Raji-19 cells, leads to the inhibition of Raji-19 cells regardless of CAR-T cell doses and dosing regimens. This is attributed to the rapid proliferation and sustained high magnitude of CAR-T cells. Conversely, when CAR-T cell proliferation rates are low and total infusion numbers are limited, a single-dose infusion provides better treatment outcomes. Our predicted bistable CAR-T cell concentration interval (ranging from 9×10^5^ to 14×10^5^ cells/mL) aligns with observed post-infusion CAR-T cell concentration intervals in clinical data (median: 480.5 cells/μL, range: 39.7−11,346 cells/μL [36]; geometric mean: 1098.79 cells/μL, range: 10^2^−10^4^ cells/μL [29]). These results collectively explain the clinical observation that a low tumour burden prior to CAR-T cell therapy is a strong positive predictor of durable response [1, 5]. Moreover, our findings emphasize that a favourable therapy outcome is heavily dependent on the interplay between CAR-T cell dose, dosing regimens, proliferation and tumour burden, which could potentially inform personalized CAR-T cell therapy strategies.

Our analysis links CAR-T cell dose, dosing regimen, and tumour burden to therapy outcomes, providing insights into clinical responses such as complete response (CR), partial response (PR), and stable disease (SD) observed in B-ALL [2, 32, 33]. We identified a critical threshold that divides the CAR-T cell concentration interval into two distinct dynamic regions. In the first interval, low tumour burdens are effectively inhibited, while high tumour burdens remain refractory. In the second interval, CAR-T cells consistently inhibit RAJI-19 cell growth.

- **Complete Response (CR)** corresponds to: (A) the first CAR-T cell concentration interval with low tumour burdens, and (B) the second CAR-T cell concentration interval with any tumour burden. In both scenarios, CAR-T cells effectively inhibit Raji-19 cell growth (Figures 4 and 5).
- **Partial Response (PR)** corresponds to a high CAR-T cell concentration within the first interval, specifically with high tumour burdens. Here, the maximal capacity of Raji-19 cell concentration decreases as CAR-T cell concentration increases, and Raji-19 cell kinetics converge to a reduced maximal capacity.
- **Stable Disease (SD)** corresponds to a high CAR-T cell concentration within the first interval, also with high tumour burdens, where CAR-T cells cannot efficiently inhibit Raji-19 cell growth.

In Section “CAR-T Cell Proliferation and Dosing Regimens Determine Raji-19 Cell Kinetics”, we proposed that single-dose infusion provides superior therapy outcomes for low proliferation rates and low infusion dosage. The CAR-T cell lysing efficiency, as described by saturated model with both RAJI-19 cell and CAR-T cell concentration, 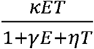, is critical. Given the rapid growth of RAJI-19 cells, the overall lysing efficiency of CAR-T cells is primarily determined by the impact of the initial dose; specifically, lysing efficiency becomes less effective as the number of RAJI-19 cells increases. With split-dose infusion, the RAJI-19 cells can proliferate faster than they are killed by the first dose, potentially leading to more target cells by the time the second dose arrives, thus rendering lysis less efficient. For high total infusion concentrations or high proliferation rates, CAR-T cell concentration is maintained at a high level or increases rapidly. Therefore, the CAR-T cell lysing efficiency remains high, and the compromised efficiency associated with the initial dose phenomenon does not occur. This finding warrants further experimental validation, particularly comparing CAR-T cells with varying proliferation capacities.

Kirouac et al utilized Hill function with respect to CAR-T cell concentration to describe CAR-T cell lysing [30]. First, using bifurcation analysis, it is easy to know that tumor kinetics (in our case, RAJI-19 cell kinetics) is only dependent on magnitude of CAR-T cells, rather than tumour burden. Moreover, we found that saturated model performs better than two semi-saturated models (purple, blue and light blue in Fig.2B). Chauhury et al [31] reviewed different CAR-T cell lysing formula by CAR-T cells, including mass action functional 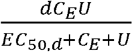. However, this formula fails to consider that tumour cell and CAR-T cell may play different roles to control saturation effect. Our model is 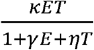, and then *γ* and *η* controls saturation with increase of RAJI-19 cell and CAR-T cell concentration, respectively. After fitting FBK assay datasets, we found that *η* (approximately 10^0.5^) is much higher than *γ* (approximately 10^-1^) among all biological replicates (Table. S4-S7, supplementary material). This indicates RAJI-19 cell concentration plays a more important role in saturation than CAR-T cell concentration does, thereby rejecting the hypothesis that tumour cells and CAR-T cells may play similar/same roles to control saturation effect. This inequivalent role of RAJI-19 cell and CAR-T cell concentration in saturation effect leads to the observed phenomenon that patients with higher tumour burdens at the start of treatment are less likely to both attain and maintain a deep response compared to those with lower tumour burden.

Flow cytometry-based killing assays are designed to quantify remaining viable RAJI-19 cell numbers following exposure to CAR-T cell therapy during a 24-72-hour incubation period. However, *in vivo* CAR-T cells typically reach a peak level within one to two weeks and then persist at a much lower level for years after infusion [11]. We suggest that CAR-T cell-induced bistable RAJI-19 cell kinetics may not be directly observed in typical FBK assays due to two potential reasons: (1) the short incubation period, and (2) the experimental designs often utilizing a fixed RAJI-19 cell number with varying CAR-T cell numbers based on E:T ratio. Firstly, our mathematical model indicates that a longer incubation period, potentially around five days, may be required for this lysis behaviours to be clearly detected (Fig. 4B and 5B). Secondly, designing FBK assays with varying Raji-19 cell numbers while maintaining a constant CAR-T cell number (or vice-versa) would increase the probability of detecting CAR-T cell-induced bistable Raji-19 cell kinetics.

One limitation of this hypothesis is that current FBK assays primarily measure RAJI-19 cells and typically do not dynamically measure CAR-T cell numbers. However, CAR-T cell proliferation is highly variable among donors, showing, for example, a 2-3-fold increase in some donors but up to a 20-fold increase in others over a seven-day culture [39]. This CAR-T cell proliferation directly influences infusion dosage, infusion frequency, and treatment outcomes. Specifically, high CAR-T cell proliferation inhibits RAJI-19 kinetics regardless of infusion cell number or single/split infusion (Figures S3 and 8). On the other hand, for low CAR-T cell proliferation, single infusion provides better treatment outcomes (Figures 7B and 7I). To overcome low CAR-T cell proliferation, increasing the infusion cell number is a feasible option. To accurately estimate infusion dosage and therapy outcomes, it is crucial to measure both RAJI-19 cell and CAR-T cell concentrations throughout the co-culture period.

The existence of CAR-T cell-induced bistable RAJI-19 cell kinetics has been predicted using RAJI-19 cell growth parameters derived from one experimental setup and CAR-T cell lysing parameters from another. To precisely identify the bistable RAJI-19 cell interval for a specific CAR-T cell, further work is required to experimentally validate this prediction within a unified experimental system. Moreover, our model predicts that with a fixed infusion dose, single-dose infusion provides superior therapy outcomes under low CAR-T cell proliferation. Future validation work should therefore include flow cytometry-based killing assays with longer incubation by different dosing regimens.

## Method

### Mathematical model

To establish a quantitative relationship between survived RAJI-19 cell and CAR-T cell concentration, we developed a family of mathematical models. These models describe different forms of CAR-T cell-mediated tumour lysis efficiency:

a. unsaturated lysing models (System 1),
b. semi-saturated lysing model with increase of RAJI-19 cell concentration (System 2),
c. semi-saturated lysing models with increase of CAR-T cell concentration (System 3),
d. saturated lysing model (System 4).

It is important to note that CAR-T cells do not immediately proliferate after simulated with RAJI-19 cells; namely, a three-day delay/lag exists for CAR-T cell proliferation after simulated (shown in Fig.1F, [37]). Because flowcytometry-based killing assay co-cultures CD19+ cell and CAR-T cell for three days, CAR-T cell concentration decrease; in this case, we utilized System 1 - System 4 to describe CAR-T cell and RAJI-19 cell kinetics over flow cytometry-based killing assay. We intended to investigate RAJI-19 cell and CAR-T cell kinetics for thirty-day co-culture. Proliferation and exhaustion of CAR-T cell is described by mathematical formular *φTE* in System 5.

Unsaturated lysing model assumes that the rate of RAJI-19 cell lysis is directly proportional to both RAJI-19 and CAR-T cell concentrations, without saturation. We describe the rate of change of RAJI-19 and CAR-T cell as 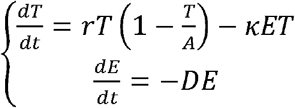 (system 1), (1) in FRA for 72-hour incubation. *T*(*t*) represents RAJI-19 cells with respect to time *t. E*(*t*) represents CAR-T cells with respect to time *t*. Parameter *κ* represents CAR-T cell lysing rate. *r* represents per capita growth rate, *r* = 0.9., *A* controls natural saturation of tumour cell replication at high tumour cell concentration, and *K* = 7.31 × 10^6^(Cells/mL/day). *D* represents natural degradation rate of CAR-T cells.

This semi-saturated lysing model incorporates saturation in the lysing efficiency as RAJI-19 cell concentration increases. We describe the rate of change of RAJI-19 cell and CAR-T cell as 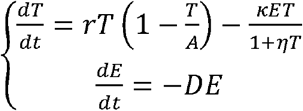 (system 2). *η* controls the saturation in lysing rate as RAJI-19 cell concentration increases.

This semi-saturated lysing model incorporates saturation in the lysing efficiency as CAR-T cell concentration increases. We describe the rate of change of RAJI-19 cell and CAR-T cell as 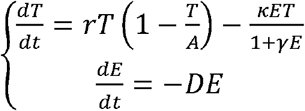, (system 3). *γ* controls the saturation in lysing rate as CAR-T concentration increases.

This saturated lysing model incorporates saturation in the lysing efficiency as RAJI-19 cell and CAR-T cell concentrations increase. We describe the rate of change of CD19+ cell and CAR-T cell as, 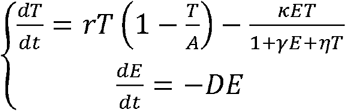 (system 4).

This saturated lysing model incorporates CAR-T cell proliferation and exhaustion. We describe the rate of change of RAJI-19 cell and CAR-T cell as 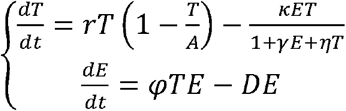 (system 5), where *φ* represents rate of proliferation and exhaustion of CAR-T cells when stimulated by RAJI-19 cells. We used *φ* = 0.3 × 10 ^−10^ day ^− 1^ cell ^− 1^ from reference [18].

### Parameter estimation

For the estimation of mathematical model parameters, RAJI-19 cell and CAR-T cell concentrations from in vitro assays were calculated. RAJI-19 cell concentration was determined using the formula: RAJI-19 concentration=5 × TN/mL, where TN represented the number of RAJI-19 cell in each well; CAR-T cell concentration was determined using the formula, CAR-T cell concentration=5 × EN/mL, where EN represented the number of CAR T cell in each well. The sum of squared error (SSE) is 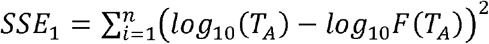, where *T*_*A*_ is experiment RAJI-19 concentration at E:T ratio of 1:1, 1:2, 1:4 and 1:8 and estimated RAJI-19 concentration at different E:T ratio. Initial guesses used for parameter estimation are *κ*_0_ = 10^−1^, *γ*_0_ = 10^−1^ and *η* = 10^−1^.

Next, model selection was performed using the Akaike Information Criterion (AIC) and the modified Akaike Information Criterion (AICc) for small sample sizes of the unsaturated, semi-saturated and saturated lysing models. The formulas were the AIC 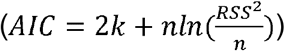 and the modified AIC 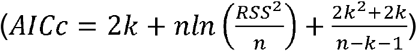 for small sample sizes of unsaturated and saturated neutralisation models were calculated, where k is the number of parameters, is the sample size, and RSS represents the residual sum of squares.

### Flow cytometry-based Killing Assay

We utilized the flow cytometry-based killing Assay data provided in Reference [28]. Briefly, non-transduced (NT) or transduced (CAR-T) effector cells were serially diluted 2-fold in a 96 well plate. RAJI-19 cell were then added to achieve effector-to-target (E:T) ratios of 1:1, 1:2, 1:4 and 1:8. Co-cultures were incubated for 72 hours in TexMACS medium supplemented with 3% human serum at 35ºC in 5% CO_2_. Cytotoxicity was determined by the number of remaining viable RAJI-19 cells by flowcytometry, with CountBright beads added to determine absolute cell numbers.

## Data Availability

https://github.com/ShlomoBergg/CAR-T-dosage-and-Raji-19

## Acknowledgements

M.L. was supported by Natural Science Foundation of China (82204265), the Science and Technology Innovation Fund of Shenzhen (JCYJ20210324101400001), Key R&D Program of Hunan Province (2023SK2082). S.X. was supported by a fellowship from Lady Davis Fellowship Trust at Technion-Israel Institute of Technology.

## Notes

### Competing Interest Statement

The authors have declared no competing interest.

### Author Declarations

https://github.com/ShlomoBergg/CAR-T-dosage-and-Raji-19, "T cells used in this study were obtained from six healthy donors and six B-cell acute lymphoblastic leukemia (B-ALL) patients." we reanalyzed the data proposed in published manuscript "AKT inhibition generates potent polyfunctional clinical grade AUTO1 CAR T-cells, enhancing function and survival" (de-identified prior to use in this study). We did not not generate any new data in our manuscript.

